# Development and Pilot testing of a Leadership Module to Support Quality Improvement Teams in Nursing Homes

**DOI:** 10.1101/2025.10.08.25336902

**Authors:** Liane R. Ginsburg, Whitney Berta, Carole A. Estabrooks, Matthias Hoben, Lonnie Kehler, Don McLeod, Jennifer Pietracci, Laurel Rose, Danielle Saj, Georgina Veldhorst, Adrian Wagg, Malcolm Doupe

**Affiliations:** School of Health Policy & Management, York University, Toronto, Ontario, Canada; Institute of Health Policy, Management & Evaluation, University of Toronto, Canada; Faculty of Nursing, University of Alberta, Edmonton, Alberta, Canada; Rady Faculty of Health Sciences, University of Manitoba, Winnipeg, Manitoba, Canada; Winnipeg Regional Health Authority, Winnipeg, Manitoba, Canada; Shared Health, Winnipeg, Manitoba, Canada; Division of Geriatric Medicine, University of Alberta, Edmonton, Alberta, Canada

**Keywords:** Implementation science, leadership, quality improvement, mixed methods, pilot, nursing homes, healthcare aides

## Abstract

**Background:** Leadership is a critical lever for supporting implementation of practice change ideas intended to improve care. We need evidence-based leadership programs to help front-line providers meaningfully implement practice change in complex care settings. Part of the SHIFT intervention, this paper describes and pilot tests a leadership program module (LeaderSHIFT) that provides training and implementation coaching to front-line leaders, as one of several integrated facilitated supports designed to help front-line care teams meaningfully enact practice change.

**Methods:** The LeaderSHIFT program module was developed based on empirical work, relevant facilitation and transformational leadership theories, and principles of stakeholder co-design and feasible engagement. A pilot implementation study was conducted that examined several of Proctor’s (2011) implementation outcomes.

**Results:** LeaderSHIFT includes four interactive workshops plus two one-on-one coaching sessions designed to develop capacity in four areas of implementation leadership: (1) *Self-awareness, (2) Motivate and inspire, (3) Facilitate learning capacity, and (4*) *Support ‘team-oriented processes’. Pilot* results suggest it can be successfully implemented (it was acceptable, adopted, appropriate, feasible). Fidelity (LeaderSHIFT role enactment) varied across pilot teams.

**Conclusions:** With a strong theoretical and empirical base, LeaderSHIFT highlights important, often overlooked, relational and socio-cultural aspects of successful implementation leadership. As such, the LeaderSHIFT program module has the potential to improve implementation of practice change interventions in nursing homes and other institutional care settings.

**Trial registration:** Registered at ClinicalTrials.gov (ID NCT03426072) on July 18, 2022.

**KEY MESSAGES REGARDING FEASIBILITY:**

1. What uncertainties existed regarding the feasibility?
  - While leadership is known to be a critical lever for implementation of evidence-informed practice change, there are few leadership training programs that have a relational focus designed to support broader team-based practice change initiatives; and uncertainty remains regarding implementability (feasibility, acceptability, appropriateness, fidelity) of this type of leadership module in complex care settings
2. What are the key feasibility findings?
  - The LeaderSHIFT module performed well on several key implementation outcomes (module acceptability, feasibility, appropriateness). Fidelity to implementation leadership was successful for managers who were able to enact relational aspects of the role.
3. What are the implications of the feasibility findings for the design of the main study?
  - Findings confirmed the value of one-to-one coaching for enhancing leaders’ relational competencies and prompted training overlap for senior and front-line leaders to ensure there is a common understanding of respective roles in intervention implementation.

## BACKGROUND

Nursing homes (NHs) are important sites of care – most nursing home residents in Canada are frail, older persons who will live out their lives in these care settings. However, while heavily relying on NHs as a care setting for older adults, societies under resource and under value nursing homes, reflecting a neglect of (or indifference to) the intersecting matters of ageing, disability, care, and care’s gendered nature (i.e., this care is mostly provided by women to women)[1]. Nursing homes have been plagued by longstanding quality concerns[2,3], worsened by COVID-19[4] and its residual effects[5,6]. Implementing interventions to bring about improvement in care processes and outcomes (hereafter referred to as ‘implementing practice change’) in these and other health settings is a necessary but immensely challenging endeavour[7]. Despite the systemic challenges, quality varies considerably between facilities within systems, suggesting that facility-level contexts and activities can make a difference[8]. Leadership is a well-known contextual antecedent for successful implementation of Quality Improvement (QI)[9,10], integrated care[11] and other evidence-based practice changes designed to improve care[12–14].

A variety of leadership training programs have been developed and used in health care settings. Many focus on developing QI leadership among clinicians or front-line teams[15–17]. Some programs that target front-line or mid-level managers are designed to develop general leadership capabilities[18], broadly support managers through organisational change[19], or enhance transformational leadership[20]. Other programs focus on promoting leadership skills specific to fostering individual-level evidence-based practice change[21–23]. However, implementation of practice change is ultimately carried out by front-line teams who provide day-to-day care. Given the non-trivial support and facilitation role that leaders play in assisting teams with implementation, practice change interventions that target front-line cross-disciplinary teams, *while also targeting leaders*, are needed[24]. We found few leadership training programs designed to support broader team-based practice change initiatives. The current paper describes the design and pilot testing of LeaderSHIFT – a leadership development module intended to help front-line leaders support cross-disciplinary teams participating in a Quality Improvement Collaborative to improve care for nursing homes residents.

### Situating LeaderSHIFT – From SCOPE to SHIFT

LeaderSHIFT is a leadership development module designed to support broader practice change interventions. It builds on a leadership training effort originally developed for the SCOPE QI intervention recently implemented in 31 Canadian nursing homes[25,26].

The ‘Safer Care for Older Persons (in residential) Environments’ (SCOPE) QI intervention is based on the Institute for Healthcare Improvement (IHI) Breakthrough Series Collaborative Model[27] and the PARIHS framework for guiding research implementation[28,29]. The aim of the SCOPE intervention was to empower front-line nursing home teams to conduct QI initiatives to improve staff work life (e.g., by fostering empowerment and building more effective care teams) and, ultimately, to improve the quality of care for residents[26,30,31]. The core components of the SCOPE intervention included (i) use of the IHI Plan-Do-Study-Act (PDSA) Model for Improvement as the core “engine for change”, (ii) quarterly in-person quality improvement (QI) learning congresses attended by all participating teams, (iii) ongoing support from an external Quality Advisor (QA) during action periods between learning congresses, and (iv) additional QA training provided to unit and facility leaders (referred to as team and senior sponsors, respectively) to help them support their frontline teams.

SCOPE teams were led by, and comprised mainly of, care aides given they provide the majority of direct care to NH residents and have intimate knowledge of residents’ daily care needs[32]. Enabling care aide leadership of the SCOPE improvement work was the primary role for team and senior sponsors. Team and Senior Sponsors in SCOPE participated in 1-hour leadership sessions at each of the learning congresses to help build their coaching and facilitation skills. Participating facilities also received a small stipend ($3000) to offset the costs of backfilling staff to participate in workshops and to cover incidental costs for supplies needed to carry out their improvement projects.

Results from SCOPE proof of principle[30], pilot[31], full trial[26], and process evaluations[25,33–35] conducted in three Canadian provinces showed that teams were committed to improving quality, that they experienced pride, empowerment, feelings of change efficacy, believed the intervention led to improvements in resident care, and they felt they forged more trusting relationships with staff in other disciples and with their managers[25,33,34]. The challenges encountered in SCOPE pertained to navigating roles, achieving buy-in from non-SCOPE colleagues, finding time to plan and carry out improvement plans, and properly incorporating more technical measurement aspects of the PDSA improvement cycle[25,34]. Despite varying levels of engagement by the team sponsor group[33], findings also showed the importance of team sponsor support for implementation fidelity[25].

To overcome the challenges just noted, and given the importance of sponsor support in SCOPE, an augmented intervention (called SHIFT - Supporting Healthcare Improvement Through Facilitation and Training) was designed that attends more purposefully to providing support to teams through a leadership module (LeaderSHIFT) that teaches unit-level leaders (team sponsors) how to more effectively assist front-line teams. SHIFT also added a readiness for change module to explicitly attend to preparing teams for the intervention (the readiness for change module is described in a companion paper).

### Objectives

As mentioned in the introduction, the key role of leadership in implementation processes is well-established. However, knowledge about ‘how’ leaders facilitate implementation of practice change in complex care settings (i.e., the process by which they do that), while growing, remains sparse. We also lack a clear understanding of how to *build* leadership support for such interventions. Building on novel work by several members of this team that explored leadership processes for facilitating implementation of SCOPE[33], the current paper has two feasibility-related[36] objectives:

1. to describe the development (theoretical foundation and design) of LeaderSHIFT (a component/module of the SHIFT intervention);
2. to present results from a small pilot implementation study of the LeaderSHIFT module to inform implementation of the full-scale SHIFT type III hybrid implementation-effectiveness trial[37].

## METHODS

To design the LeaderSHIFT module, we drew on empirical work regarding the processes by which leadership influences implementation of a QI practice change intervention, relevant facilitation and transformational leadership theories, and we employed principles of stakeholder co-design and feasible engagement (these procedures for module design are described as part of the Methods section; the module itself is described in the Results section). The LeaderSHIFT module underwent an implementation pilot[38] with a concurrent process evaluation. This study adheres to relevant sections of the Template for Intervention Description and Replication (TIDierR)[39] and CONSORT (Consolidated Standards of Reporting Trials) extension for pilot and feasibility trials[40].

### Defining LeaderSHIFT

The LeaderSHIFT module was designed using results of the SCOPE process evaluation regarding ‘how’ teams implemented SCOPE[25] and the specific processes by which front-line and senior leaders (team and senior sponsors, respectively) facilitated implementation of SCOPE[33]. These findings were used to define the core elements of LeaderSHIFT to ensure the module reflected empirical work as well as relevant theory regarding facilitation[29,41–43], and transformational leadership [44–47]. Most empirical work in the field of implementation science identifies *determinants* of implementation success without illuminating the more dynamic *processes* and embedded mechanisms of implementation. A strength of LeaderSHIFT is that it is designed using the relatively small body of empirical work that does exist regarding the *processes/mechanisms* by which leaders have an impact on implementation.

In keeping with the principle of feasible engagement central to the Knowledge-to-Action (KTA) framework[48], LeaderSHIFT was also designed to be consistent with health system priorities and initiatives. As such, LeaderSHIFT aligns with the LEADS for a Caring Environment framework[49]. The LEADS framework, which is “a statement of aspirations for the practice of leadership”[50], is part of the health system’s approach to leadership development in Manitoba where SHIFT is being implemented. In addition, recent research in Canadian nursing homes suggests leadership practices that create conditions for high-quality LTC are consistent with capabilities outlined in the LEADS framework[51].

#### User Involvement

Elements of co-design and integrated knowledge translation (iKT) were included in the design of LeaderSHIFT by soliciting perspectives of key stakeholders including intervention targets (leaders) through a pilot study (described below). Perspectives of system-level decision makers who would be responsible for scale up of the SHIFT program in Manitoba, including the LeaderSHIFT module, were obtained through their involvement on a Study Operations Group which met on an ad hoc basis to consider feasibility issues and to ensure SHIFT / LeaderSHIFT fit with health system priorities. The final LeaderSHIFT module, including the ways in which theories, frameworks and prior empirical work informed the program, is described in the results section.

### Pilot Testing LeaderSHIFT

The LeaderSHIFT module (described in the results section) was pilot tested as part of the SHIFT pilot, with three units in two nursing homes in the Canadian province of Manitoba in 2023 (convenience sample). A mixed-methods concurrent process evaluation was used to evaluate the SHIFT pilot, including the LeaderSHIFT module. Using methods and tools previously employed by members of this team[25], a combination of Quality Advisor diaries, observer field notes, team sponsor surveys, and focus groups were used to evaluate LeaderSHIFT in the pilot. Table 1 summarizes these data collection approaches.

**Table 1:**
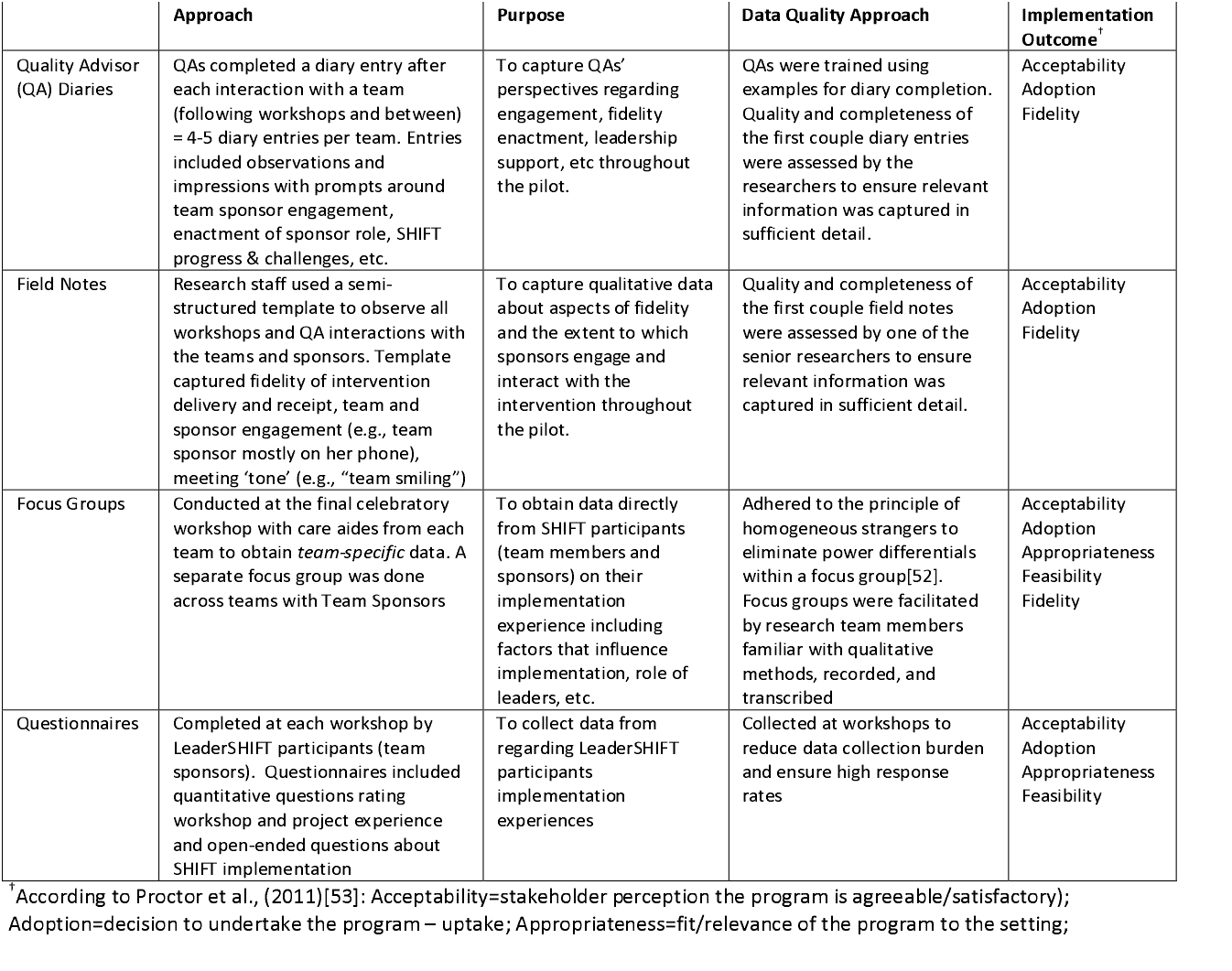
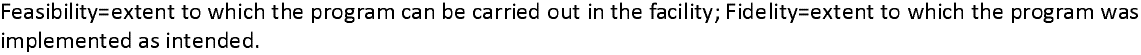
Pilot Data Collection Approaches & Implementation Outcomes Relevant to LeaderSHIFT.

|  | Approach | Purpose | Data Quality Approach | Implementation Outcome<sup>†</sup> |
| --- | --- | --- | --- | --- |
| Quality Advisor (QA) Diaries | QAs completed a diary entry after each interaction with a team (following workshops and between) = 4-5 diary entries per team. Entries included observations and impressions with prompts around team sponsor engagement, enactment of sponsor role, SHIFT progress & challenges, etc. | To capture QAs' perspectives regarding engagement, fidelity enactment, leadership support, etc throughout the pilot. | QAs were trained using examples for diary completion. Quality and completeness of the first couple diary entries were assessed by the researchers to ensure relevant information was captured in sufficient detail. | Acceptability<br>Adoption<br>Fidelity |
| Field Notes | Research staff used a semi-structured template to observe all workshops and QA interactions with the teams and sponsors. Template captured fidelity of intervention delivery and receipt, team and sponsor engagement (e.g., team sponsor mostly on her phone), meeting 'tone' (e.g., "team smiling") | To capture qualitative data about aspects of fidelity and the extent to which sponsors engage and interact with the intervention throughout the pilot. | Quality and completeness of the first couple field notes were assessed by one of the senior researchers to ensure relevant information was captured in sufficient detail. | Acceptability<br>Adoption<br>Fidelity |
| Focus Groups | Conducted at the final celebratory workshop with care aides from each team to obtain *team-specific* data. A separate focus group was done across teams with Team Sponsors | To obtain data directly from SHIFT participants (team members and sponsors) on their implementation experience including factors that influence implementation, role of leaders, etc. | Adhered to the principle of homogeneous strangers to eliminate power differentials within a focus group[52]. Focus groups were facilitated by research team members familiar with qualitative methods, recorded, and transcribed | Acceptability<br>Adoption<br>Appropriateness<br>Feasibility<br>Fidelity |
| Questionnaires | Completed at each workshop by LeaderSHIFT participants (team sponsors). Questionnaires included quantitative questions rating workshop and project experience and open-ended questions about SHIFT implementation | To collect data from regarding LeaderSHIFT participants implementation experiences | Collected at workshops to reduce data collection burden and ensure high response rates | Acceptability<br>Adoption<br>Appropriateness<br>Feasibility |

<sup>†</sup> According to Proctor et al., (2011)[53]: Acceptability=stakeholder perception the program is agreeable/satisfactory); Adoption=decision to undertake the program – uptake; Appropriateness=fit/relevance of the program to the setting;
Feasibility=extent to which the program can be carried out in the facility; Fidelity=extent to which the program was implemented as intended.

#### Pilot Outcomes & Analysis

*Five of* the eight implementation outcomes defined by Proctor et al., (2011) that are relevant to a pilot study (e.g., penetration and sustainability could not be assessed in this pilot context) were examined to explore the acceptability, adoption, appropriateness, feasibility, and fidelity of the LeaderSHIFT module. *Adoption* was assessed at the end of each LeaderSHIFT workshop with an exit survey question about whether participants would be able to use the information shared. *Feasibility* was assessed with an item asking Team Sponsors whether the responsibilities being asked of them were manageable (5-point Likert-type agreement scales were used for both items). All five implementation outcomes were also assessed using qualitative data described in table 1. Pilot data were content analyzed by two of the authors (LG and LK) to explore the five implementation outcomes noted above and to suggest modifications to the LeaderSHIFT module. A final LeaderSHIFT module was defined.

## RESULTS

### LeaderSHIFT Module - Focus and Topics

Consistent with LEADS and SCOPE results, LeaderSHIFT is predicated on the idea that “leadership is best understood in the context of both human and mechanistic systems”[50], although SCOPE results suggest considerable emphasis should be placed on the human systems piece[33]. At its core is the recognition that, to support practice change, leaders need to move beyond transactional behaviours and removal of barriers. Leaders need to engage in social / relational processes that inspire and build a common vision, and that build mutually supportive relationships and interactive problem-solving approaches characteristic of higher-order learning[33,41,42,54]. LeaderSHIFT emphasizes the interpersonal, relational, and socio-cultural aspects of leadership while also recognizing the need for leaders to provide material support.

Guided by the LEADS framework and findings from the SCOPE process evaluation and analysis of leadership practices in SCOPE, LeaderSHIFT focuses on developing capacity in four areas to promote implementation leadership: (1) *Self-awareness*, reflection, and personal growth to ensure the leader is developing and is positioned to lead, to share leadership, and to support others to play leadership roles in the practice change; (2) *Motivate and inspire* by demonstrating their own commitment to the change initiative and making clear their role in supporting teams to maintain their commitment; (3) *Facilitate learning capacity* within teams by fostering team member participation in the change initiative and empowering team members to step into team leadership roles; (4) *Support ‘team-oriented processes’* that develop and nurture relationships with front-line team members and that support team members as they navigate relationships with each other and other staff to implement practice change.

Table 2 provides a matrix outlining the LeaderSHIFT module topics, a sample of corresponding activities, and shows how each topic maps to SCOPE results (which are themselves based on relevant leadership and facilitation theories) and aspects of the LEADS framework. The LEADS framework and the SCOPE results complemented each other in several important ways to guide development of four LeaderSHIFT sessions. First, implementation leadership in healthcare is about doing the work in a complex system. LEADS’ System Transformation capabilities around encouraging and supporting innovation and orchestrating change are central to SCOPE’s focus on quality improvement in complex systems and are relevant to the LeaderSHIFT **Orientation** (session 1) and session 2, **Leading in a Complex System**.

**Table 2:** LeaderSHIFT Topic Matrix: Map to Theoretical Frameworks and Empirical Work.

| Area of Capacity Development | LeaderSHIFT Module Elements | Sample Workshop Activities / Questions | Rooted in Transformational Leadership[44,45] and Facilitation Theory[29,42] |  | LEADS Framework & Relevant Capabilities[49]<br>Canadian College of Health Leaders |
| --- | --- | --- | --- | --- | --- |
|  |  |  | SCOPE Program Theory<br>*Ginsburg et al., [25]* | How Leadership Influences Implementation<br>*Ginsburg et al., [33]* |  |
| Role Clarity | **Orientation (LS1)**<br>Virtual session with Senior & Team Sponsors<br>Review of roles and commitments<br>Recruiting and supporting SHIFT teams<br>Overview of core leadership capabilities as laid out in LEADS<br>Overview of SHIFT initiative | Discussion areas:<br>What is the difference between authority and leadership?<br>Who will you recruit for your team and how will you ensure team members have *time (weekly meetings)* and *support* to lead change activities?<br>Complete LEADS Capabilities Self-Assessment | All 4 SCOPE implementation processes speak to leaders' roles, and how to build teams that can engage in these processes:<br>Learning and Applying QI Knowledge and Skills (P1)<br>Negotiating (developing/fostering) relationships (P2)<br>Navigating workplaces (P3)<br>Shifting culture around roles / hierarchy (P4) | All 3 SCOPE leadership processes speak to leaders' roles:<br>Develop and Affirm Commitment (T1)<br>Facilitate Learning Capacity (T2)<br>Support Team-oriented Processes (T3) |  |
| Promoting System Integration through Leadership Roles | **Topic 1 – Leading in a Complex System (LS2)**<br>Virtual session with Team Sponsors<br>Leading change in a Complex System (volatile, uncertain, complex, ambiguous)<br>Adaptive leadership in complex systems<br>Shifting the role of the Leader in supporting change (distributed vs traditional leadership)<br>Managing the human side of change – Introduction to topics 2 & 3 | Reflection exercise re 4 leader-relevant Program Theory processes (human side of change):<br>1. Develop supportive learning environment<br>2. Foster positive team dynamics<br>3. Build resilience against challenges<br>4. Enhance empowerment, build trust | Teams navigate workplaces by finding capacity for SCOPE (P3a) and building team resilience against challenges associated with competing (often higher) priorities (P3b).<br>Teams build resilience against challenges with the PDSA material and doing the associated improvement work; sponsors support teams even when they experienced failure (P3b). | Sponsors bridge SCOPE with facility context: Engage and support teams within current context (leveraging current priorities / previous training; working within organizational pressures (T1b) | **System Transformation**<br>**Capabilities:**<br>Demonstrate systems/critical thinking<br>Encourage and support innovation<br>Orient themselves strategically to the future<br>Champion and orchestrate change |
| **One-to-one Coaching Session 1** (pre-LS3)<br>Emphasis on 'Lead Self'<br>Discuss the core capabilities of an adaptive / agile leader that are needed to support teams to make change in complex systems |  |  |  |  |  |
| Self-awareness<br>Motivate and Inspire | **Topic 2 – Lead (Develop) Self & Inspire Vision (LS3)**<br>In-Person session with Team Sponsors<br>Reflect on and identify perceptions and assumptions about change and change leadership<br>Take responsibility for managing emotions, mindsets, including in the face of unexpected events<br>Seek out opportunities for learning and development<br>Recognize and demonstrate character traits required to inspire change<br>Use various communication strategies to ensure the team is committed to a shared purpose and is clear about their improvement goals | Review of LEADS Capabilities Self-Assessment<br>Developing an achievable Personal Learning Plan<br>Group discussion/check in re personal learning goals<br>Above/Below the Line<br>Exercise to create awareness of how you engage your team (and what might be getting in the way):<br>how you respond to barriers that arise in the change process<br>What are the implications? | Sponsors develop a more distributed approach to leading to promote a supportive learning environment (P1b)<br>Sponsor perceptions of front-line teams' abilities and capacity to lead change shifted (P4a) | Sponsors developed commitment to the intervention and their leadership role in it, and demonstrated this by inspiring team members (T1a) and tending to supportive (pragmatic) aspects of the role (T1b)<br>Mutual trust between sponsors and followers developed over time as sponsors experienced growth, expressed confidence in and respect for their followers' abilities (T3a) | **Lead Self**<br>**Capabilities:**<br>Are self-aware<br>Manage themselves<br>Develop themselves<br>Demonstrate character<br>**Achieve Results (inspiring vision)**<br>**Capabilities:**<br>Set direction (by inspiring clear vision)<br>Strategically align decisions with vision, values, and evidence<br>Assess and evaluate |
| **One-to-one Coaching Session 2** (pre-LS4)<br>Emphasis on 'Engage Others'<br>Discuss the core capabilities of an adaptive / agile leader that are needed to support teams to make change in complex systems |  |  |  |  |  |
| Facilitate learning capacity<br>Support team-oriented processes | **Topic 3 – (Tangible) Engagement of Others & Coalition Building (LS4)**<br>In-Person session with Team Sponsors<br>Recognize the work and effort of team members during the | Balcony-Dance Floor exercise<br>Invite participants to reflect and share what it feels like to be on the "dance floor" and what it feels like to be on the balcony observing themselves and their | Sponsors develop an environment to support team learning about improvement work (P1b)<br>Sponsors enable empowerment of teams and development of trust between Sponsor and | Sponsors engage in strategies to increase participation and promote critical reflection; created environments where followers felt safe to actively engage with | **Engage Others**<br>**Capabilities:**<br>Foster the development of others<br>Contribute to creation of healthy |
|  | improvement process<br>• Create and foster trust and interdependence in relationships among team members<br>• Create collaborative opportunities for the team to learn and utilize each team member's strengths<br>• Distributed leadership and empowerment<br>• Navigating relationships with those external to the team | team.<br>Reflection exercise “Living in a perfect storm”<br>• Reflections on challenges of leading change in a complex system | teams’ to enable teams to lead change (P4a)<br>• Sponsors help the team attain legitimacy on the unit by leveraging their position to ease resistance (P2b) | SCOPE activities (T2a)<br>• Sponsors identify and support opportunities for care aide leadership (T2b)<br>• Sponsors develop and nurture relationships with their followers which built mutual trust and high levels of team engagement (T3a)<br>• Sponsors navigate relationships with other staff - removing external relational barriers to allow team to move forward (T3b) | organization<br>• Communicate effectively<br>• Build teams<br>***Develop Coalitions***<br>**Capabilities:**<br>• Purposively build partnerships and networks to create results<br>• Demonstrate a commitment to service<br>• Navigate socio-political environments |

Second, the focus on self-development and growth, modeling character, and being accountable from LEADS ‘Lead Self’ dimension was a more passive theme in the SCOPE results – there was evidence of this type of personal growth, but it is more explicit and strongly emphasized in the LEADS framework. Developing capacity in this area is included as part of session 3, **Lead / Develop Self & Inspire Vision**.

Third, the ‘Engage Others’ LEADS dimension has strong overlap with SCOPE results regarding successful processes for engaging teams in practice change implementation; however, it has less emphasis on inspiring others to share the vision and mission of the change effort as they engage others. Evidence of this key inspirational aspect of transformational leadership was strong in the SCOPE results. Although ‘Inspiring vision’ is a capability in LEADS’ ‘Achieve Results’ domain, the description (setting direction and establishing and communicating expectations) suggests a more authoritative than distributed approach to achieving results – something that may be more appropriate for a general leadership framework like LEADS than for LeaderSHIFT which is about enabling front-line leadership to support implementation of practice change. LeaderSHIFT session 3 is therefore devoted to **Lead / Develop Self & Inspire Vision**.

Finally, the ‘Navigate socio-political environments’ capability in LEADS’ ‘Develop Coalitions’ dimension aligns with processes leaders in SCOPE employed to engage staff outside the SCOPE team and supports LeaderSHIFT session 4 which is devoted to **Engagement of Others & Coalition Building**. Sessions 1 and 2 establish expectations and set the stage while sessions 3 and 4 seek to develop capacity in the four areas (noted above) designed to strengthen leadership support for practice change implementation: *Self-awareness, Motivate and inspire, Facilitate learning capacity*, and *Support ‘team-oriented processes’*. All four LeaderSHIFT sessions are delivered by an external Quality Advisor (QA) with expertise in facilitation and prior NH leadership experience. Session 1-2 are delivered virtually, sessions 3-4 are delivered face-to-face in off-site meeting space.

### Pilot Results

Three team sponsors participated in the LeaderSHIFT module pilot (one from each of the three units that participated in the SHIFT pilot). Ten healthcare aide team members participated in the SHIFT pilot (3-4 from each of the three teams). Survey results reported below are based on data from the team sponsors. Results regarding enactment of the LeaderSHIFT role draw on data from two focus groups, one conducted with team sponsors and one conducted with team members.

#### LeaderSHIFT Implementation

Several outcomes from Proctor et al’s., taxonomy of implementation outcomes[53] suggest LeaderSHIFT was implemented successfully. Results support *acceptability* (team sponsors strongly agreed the LeaderSHIFT sessions were valuable, one participant stated “It’s all indeed valuable and informative”), *adoption* (sponsors strongly agreed they would be able to use the information shared), *feasibility* (sponsors agreed or strongly agreed the responsibilities being asked of them were manageable, and participants attended all delivered LeaderSHIFT sessions), and *appropriateness* (one respondent stated “The topic was relevant and useful).

Using Bellg’s three aspects of *fidelity*, field notes and surveys respectively indicate the LeaderSHIFT module was delivered as intended, and concepts were understood by participants. Focus group results from team members, diaries, and field notes suggest team sponsors in two of the three participating units were able to enact their internal facilitation role and support their teams with fidelity (i.e., in the manner intended by LeaderSHIFT). One team sponsor’s comment embodies both the intrinsic motivational aspect and the importance of support to team sponsors from the senior sponsor. When asked what else, other than funds, is needed to help ensure that team sponsors feel supported to participate in SHIFT, one Team Sponsor said, “It’s participating and engaging and moving forward with learning opportunities for quality improvement…for me that’s enough as a reward. If the money [facility stipend] is there, then that’s a bonus. And it doesn’t come from my cost centre…and, you know, having your direct report be respectful of what you’re doing” (TS3).

The team sponsor who did not successfully enact their role was simply not engaged. The final QA diary entry for this team noted that “the team expectation of the TS is low to none so when the team got stuck, no support was given. No dedicated time allotted”. In their team member focus group, when asked about support received from their team sponsor, one respondent said “no, nothing”, another said “No. We even said when we were driving here, why is she coming, because she wasn’t a part of anything” (TM FG, site 3).

#### LeaderSHIFT Module Design and Topics

During the SHIFT pilot, the Senior Sponsor (usually a facility’s Director of Care) was targeted for facility recruitment into the SHIFT program but was not formally included in LeaderSHIFT after that point (LeaderSHIFT focused on Team Sponsors – normally unit level managers). Pilot field notes and focus group data showed very limited communication between the Senior and Team Sponsors (e.g., two of three team sponsors were not aware there was a facility stipend available to offset minor costs associated with participation in SHIFT). This finding suggested the team and senior sponsors should participate together in the first LeaderSHIFT session to enhance role clarity and to ensure leaders at both levels receive a common orientation to the project. When asked if the senior sponsor should attend additional learning sessions, team sponsors agreed some overlap early on would help but “Not all the time, right…maybe just the first – like the initial meeting, yeah [TS1]…to demonstrate support [TS3]”.

Pilot focus group data also suggested it would be useful to provide additional guidance to sponsors regarding key operational issues pertaining to how best to identify team members (a common suggestion was choosing care aides “with the most influence” [TS1]) and the importance of enabling regular team meetings during action periods between learning congresses. Finally, the pilot suggested that, to facilitate team sponsor engagement and to enhance their personal growth and capacity to support teams, team sponsors participating in LeaderSHIFT would benefit from the addition of one-on-one coaching. Importantly, the pilot did not suggest changes to LeaderSHIFT topics/emphasis, duration, or intensity.

#### Final LeaderSHIFT Structure

The LeaderSHIFT module is designed to function as part of, and parallel to, a larger practice change initiative – in this case LeaderSHIFT is one of three modules in the SHIFT QI program which is intended to train front-line teams to implement change. The broader SHIFT program is delivered over an 8-month period and begins by engaging senior leaders in readiness for change activities (module 1). It then engages senior and (primarily) team sponsors through the LeaderSHIFT module (module 2). Based on the pilot results and final consultation with our system-level decision maker group, the final LeaderSHIFT module was structured as follows.

- Senior and team sponsors participate together in the initial LeaderSHIFT learning session 1 (LS1 - orientation) to ensure a common understanding of SHIFT, available resources (i.e., the stipend) and respective roles.
- Team sponsors receive training through three additional collaborative learning sessions (LS2-4) (outlined in table 2).
- Team sponsors take part in two 45-minute one-to-one coaching sessions, prior to learning sessions three and four, conducted by the external facilitator who delivers LeaderSHIFT.
- Team and Senior Sponsors are encouraged to attend the broader SHIFT Learning congresses where teams learn about and work on PDSA cycles (SHIFT module 3). For purposes of
- logistical efficiency, LeaderSHIFT sessions 3 & 4 take place immediately following SHIFT learning congresses.
- All four LeaderSHIFT learning sessions should be 1.5 to 2 hours to allow enough time for content discussion and reflection.

Figure 1 shows the structure of the final SHIFT intervention, with LeaderSHIFT (module 2) highlighted. The full SHIFT program and the readiness for change module are described in two other manuscripts currently under review.

**Figure 1:**
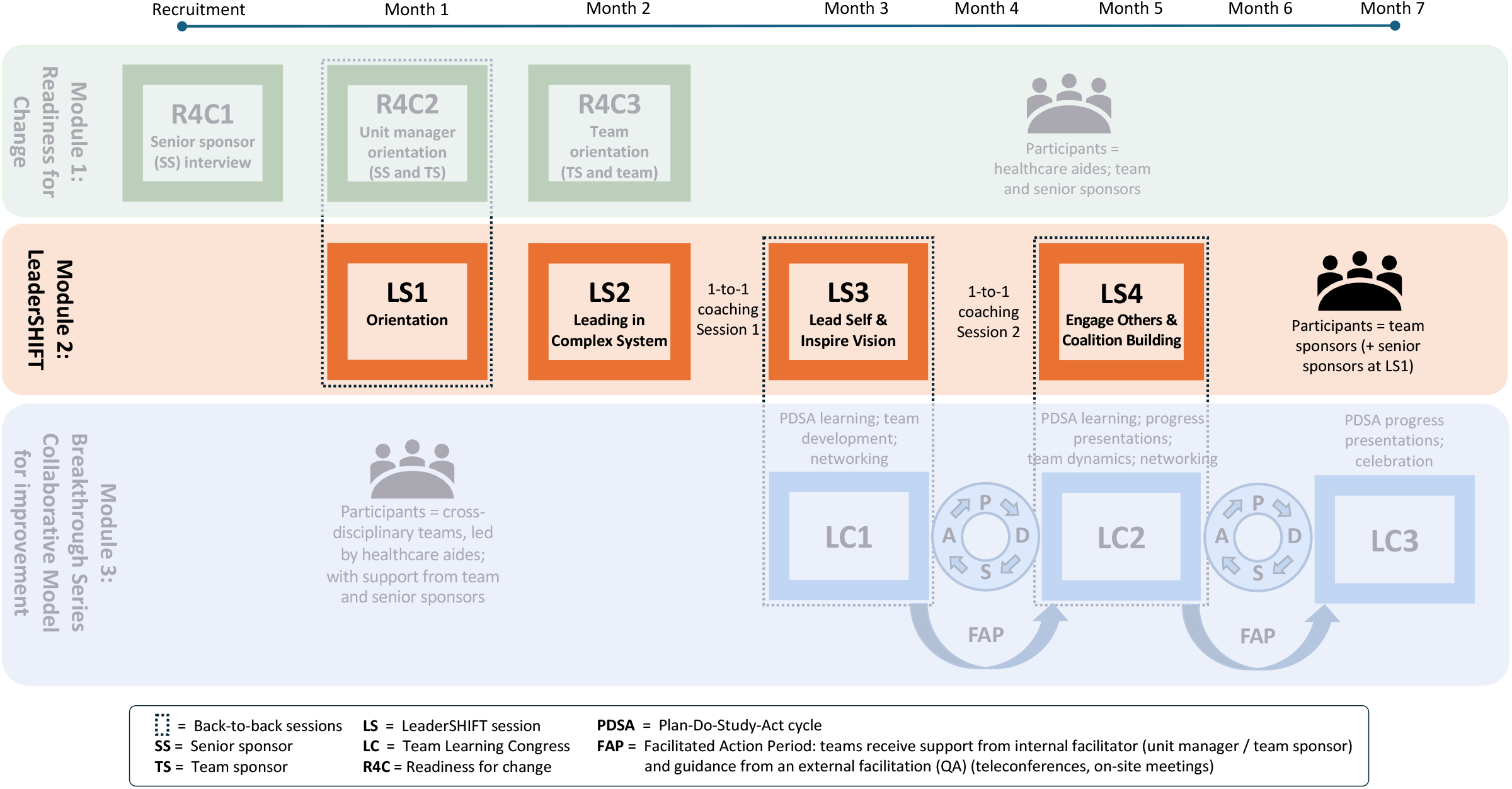
SHIFT Program Structure.

## DISCUSSION

We describe the development and piloting of LeaderSHIFT – a module designed to help engage leaders to support front-line led, cross-disciplinary teams participating in a Quality Improvement Collaborative designed to improve care for nursing home residents. Our prior work showed that leadership support is a key determinant of intervention implementation[25]; we also described processes by which leaders, with minimal additional training, support implementation[25,33]. Building on this work, the current paper describes the design and piloting of an enhanced leadership module intended to further strengthen leadership capacity to support implementation of practice change initiatives. Pilot results suggest LeaderSHIFT can be successfully implemented and that participation in LeaderSHIFT is not overly burdensome.

LeaderSHIFT aligns with a limited body of empirical work [51,55,56] on the processes / mechanisms by which leaders can support and engage staff to implement change. This evidence demonstrates (1) that transformational and other forms of positive leadership which are central to LeaderSHIFT (and which emphasize a moral perspective, role-modelling, follower self-determination, and positive social exchanges with followers) have a positive effect on staff work engagement[47,51,52], and (2) that mutual trust, follower autonomy and competence are important mechanisms that mediate the relationship between leadership and staff engagement in practice change interventions[56].

Importantly, the LeaderSHIFT module addresses leader “roles” and “feasible engagement” with those roles. The module provides dedicated support, teaching, and individual coaching for leaders. Leaders develop themselves and learn to engage and inspire their teams. Recognition regarding the value of role development was evident in a recent study of a program to develop leadership for evidence-based practice implementation[57]. In that study, participants noted that learning about their own leadership behaviours and about implementation broadly—what is required and the nature of their role in supporting change— had more lasting benefits than typical leadership training which focuses on developing particular skills associated with a new practice[53]. Emphasizing role development over skill content is also consistent with a recent call for leadership development programs to emphasize the system context and encourage holistic approaches to leadership development that foster ongoing self-authored growth over periodic program-based training[58]. The idea is that role development is more likely to lead to sustainment of implementation leadership practices.

It is important to acknowledge that the impact of leadership training programs can be limited by the organizational context and culture within which leaders function[59]. However, given that leadership is the primary culture embedding mechanism[60], we and others[61] contend that the centrality of role development in LeaderSHIFT, with its emphasis on growth, engagement, and system transformation and improvement, should promote organizational cultures that are conducive to change.

LeaderSHIFT shares certain properties found in Leadership and Organizational Change for Implementation (LOCI) which is a multi-pronged implementation strategy designed to enhance both transformational and transactional leadership and improve implementation leadership and climate within organizations[23]. Like LeaderSHIFT, LOCI targets front-line managers, includes training and individual coaching, but also recognizes the importance of building senior leadership support and, ultimately, the role of leaders in shaping a climate conducive to change. Where LeaderSHIFT differs from LOCI is in its’ focus on interventions that target practice change carried out by teams rather than individual providers, and in the intensity of the program. While the SCOPE program which SHIFT was adapted from was similar in intensity to LOCI (6-7 days of training over 12 months), LeaderSHIFT was reduced (to promote feasible engagement) and includes fewer training days spanning a shorter period (8 months). Coaching in LeaderSHIFT occurs twice over the 8-month period while LOCI coaching takes place monthly. A lower intensity program is supported by SCOPE[25] and LOCI[62] evaluations showing time was among the highest managerial barriers to implementation as well as a meta-analysis showing the number of coaching sessions is not a significant moderator of coaching’s positive effects[63].

The one-to-one coaching that is part of LeaderSHIFT is not typical of leadership training programs. Coaching targets individuals and the process of helping them gain insights to actualize their own potential and outcomes around goal attainment[64]. A recent systematic review in healthcare[65] and a broader meta-analysis[63] both support the positive effects of stand-alone coaching on leadership development. Other studies have found stand-alone coaching programs targeting front-line and mid-level managers have a positive impact on developing leadership for organizational change[66,67]. A key mechanism of impact in these and other studies is consistent with the focus of leader development in LeaderSHIFT and suggests coaching has its impact, at least in part, through improvements in leaders’ perspective-taking and self-insight[66], and improvements in ‘authentic’ leadership and establishing meaningful relationships[68].

The approach in LeaderSHIFT of using coaching, not as a stand-alone program, but as part of a larger leadership training / development module was seen in a smaller number of studies[69–71]. However, owing to our prior detailed process evaluation findings, LeaderSHIFT is somewhat unique in its heavy emphasis on relationship building between leaders and followers as a mechanism to support team-based implementation of practice change. In LeaderSHIFT, coaching is not only a promising avenue for promoting greater personal growth among engaged leaders, but it has considerable value in terms of retaining leaders who, for individual or contextual reasons, may be less engaged in the change initiative (as was evident for one of the three leaders in our pilot). Coaching within the LOCI intervention served a similar ‘continuous engagement’ function[57].

Designed for feasible engagement, LeaderSHIFT uses virtual sessions for initial orientation work and within-team relationship building (i.e., between levels of leadership within an organization) plus face-to-face collaborative learning experiences that engage leaders across organizations. The emphasis on roles rather than content and the fact that the LeaderSHIFT module takes place within the context of a learning collaborative with leaders from multiple facilities positions the module well for promoting learning from a complexity science perspective. Collaborative learning sessions provide peer support[57] and are vehicles for interrelating and reflection, which are themselves important approaches for managing change in complex systems[72].

The structure of the LeaderSHIFT module is also useful for addressing practical barriers to implementing QI and other complex changes. Leader engagement, emphasizing the leader’s role, is front-loaded prior to engaging the team while later LeaderSHIFT sessions that take place during teams’ improvement work address practical challenges leaders and teams struggle with such as engaging staff beyond the project team.

Pilot results support proceeding with testing the LeaderSHIFT module as part of the full scale SHIFT type III hybrid implementation-effectiveness trial. The full SHIFT trial results will explore the role of LeaderSHIFT in the broader implementation of SHIFT, including results regarding (1) the extent and ways in which team sponsors are able to enact LeaderSHIFT approaches and roles described in this paper, and (2) the extent and tangible ways leadership support facilitates other aspects of SHIFT implementation and SHIFT outcomes.

## CONCLUSIONS

The current paper describes the design and piloting of an enhanced leadership module intended to strengthen leadership capacity to support implementation of QI and other practice change initiatives. LeaderSHIFT aligns with an emerging body of empirical work on the processes / mechanisms by which leaders can support and engage staff to implement change. LeaderSHIFT addresses leader “roles” and “feasible engagement” with those roles. The module provides dedicated support, teaching, and individual coaching for leaders. Leaders develop themselves and learn to engage and inspire their teams. Pilot results suggested minor modifications to the LeaderSHIFT program module and indicate it can be successfully implemented and that participation is both feasible and acceptable to participants.

## Data Availability

Data generated and/or analysed during the current study are not publicly available in accordance with the privacy and confidentiality requirements of the University of Manitoba Research Ethics Board. Select data may be available from the corresponding author on reasonable request.

## LIST OF ABBREVIATIONS

FAP: Facilitated Action Period
IHI: Institute for Healthcare Improvement
TM: Team member
TS: Team sponsor (unit manager)
NH: Nursing Home
PDSA: Plan-Do-Study-Act cycle
QA: Quality Advisor
QI: Quality Improvement
R4C: Readiness for Change
SCOPE: ‘**S**afer **C**are for **O**lder **P**ersons (in residential) **E**nvironments’ (Intervention)
SHIFT: **S**upporting **H**ealthcare Improvement Through **F**acilitation and **T**raining (Intervention)
SS: Senior Sponsor (facility senior leader)

## DECLARATIONS

### Ethics approval and consent to participate

This study was reviewed and approved by the University of Manitoba (Bannatyne Campus) Health Research Ethics Board (HREB), reference number: HS18486 (H2015:045). Written consent was obtained from all study participants.

### Consent for publication

N/A

### Competing interests

The authors declare that they have no competing interests

### Funding

SHIFT is part of the larger Supporting Older Adult Healthcare Reform through Research (SOARR) research program, funded by Manitoba Health, Seniors and Long-Term Care (MHSLTC) through a Service Purchase Agreement between the Government of Manitoba and the University of Manitoba. The results of this study are those of the authors, and no official endorsement by MHSLTC is intended or should be inferred.

### Authors’ contributions

LG, DM and MBD lead the development of the LeaderSHIFT module. LR and GV contributed to design feasibility. MBD, LG, WB, MH, CE and AW conceived and refined SHIFT design and measurement. LK, JP, and DS operationalized all day-to-day aspects of LeaderSHIFT, including data collection. LG prepared the initial draft of this manuscript, and all authors reviewed and approved the final manuscript version.

## Acknowledgements

We wish to thank Cristie Perfas for her work and dedication to the QI teams and their team sponsors. We are also grateful to the organizations and care teams who participated in and enabled this study, particularly the team sponsors for their engagement and feedback.

## Notes

### Competing Interest Statement

The authors have declared no competing interest.

### Clinical Trial

NCT03426072

### Author Declarations

This study was reviewed and approved by Health Research Ethics Board (HREB) of the University of Manitoba (Bannatyne Campus) who gave ethical approval for this work (Ref number: HS18486 (H2015:045)).

